# Factors associated with SARS-CoV-2 among persons living with HIV in Zambia: Analysis of three concurrent SARS-CoV-2 prevalence surveys conducted in July 2020 in six districts of Zambia

**DOI:** 10.1101/2023.10.10.23296849

**Authors:** Stephen Longa Chanda, Cephas Sialubanje, Mukumbuta Nawa, Nyambe Sinyange, Warren Malambo, James Zulu, Dabwitso Banda, Paul Zulu, Jonas Hines

**Affiliations:** Zambia National Public Health Institute, Surveillance and Disease Intelligence Cluster, Lusaka Zambia; School of Public Health, Department of Environmental and Public Health, Levy Mwanawasa Medical University, Lusaka, Zambia; United States Centers for Disease Control and Prevention, Lusaka, Zambia; Ministry of Health, Lusaka, Zambia

**Keywords:** HIV infections, SARS-CoV-2, prevalence, reverse transcriptase polymerase chain reaction, enzyme-linked immunosorbent assay, Zambia, COVID-19 vaccines

## Abstract

People living with HIV (PLHIV) are a unique population because of their altered immune systems and taking antiretroviral therapy (ART) that may modify risk of SARS-CoV-2. Evidence from sub-Saharan African countries suggests that, despite not having higher SARS-CoV-2 infection prevalence than HIV-negative persons, PLHIV suffer worse COVID-19 outcomes. We assessed the prevalence of SARS-COV-2 infection by HIV status in Zambia in July 2020.

We analyzed data from three different concurrent SARS-CoV-2 prevalence surveys (household, outpatient-department, and health-worker) conducted in six districts of Zambia in July 2020. Information on demographics and medical history was collected. Nasopharyngeal swabs were used to screen for SARS-CoV-2 RNA using polymerase-chain-reaction (PCR) and blood specimens were screened for SARS-CoV-2 virus-specific antibodies using an enzyme-linked-immunosorbent-assay (ELISA). Test-specific SARS-CoV-2 prevalence was calculated. Multilevel logistic regression models were used to measure test-specific adjusted odd ratios (aORs) of SARS-CoV-2 positivity by HIV status, controlling for demographic and medical history.

We analysed the outcomes of the two different tests separately. Among 7,092 participants, 4,717 (66.5%) consented to blood-draw and 4,642 (65.5%) consented to nasopharyngeal swab. Overall SARS-CoV-2 positivity was 9.4% by PCR and 3.8% by ELISA. SARS-CoV-2 prevalence detected by PCR was higher among PLHIV than HIV-negative respondents (12.4% vs 9.1%, respectively, OR: 1.4, 95% confidence intervals [CI]: 1.0-1.9) and lower by ELISA (1.9% vs 3.9%, respectively, OR: 0.5, 95%CI: 0.2-0.9). Among PLHIV, not being on ART was an independent predictor of SARS-CoV-2 PCR positivity (aOR: 5.24,95% CI: 1.19-22.22) but did not have a significant effect on ELISA results.

During the first COVID-19 wave in Zambia, PLHIV were more likely to be acutely infected with SARS-CoV-2 but less likely to be seropositive than participants without HIV. Intervention programs could focus on early access to COVID-19 vaccinations, testing and ART might reduce COVID-19 morbidity among PLHIV.

## Introduction

From March 2020 through January 2023, over 339,000 laboratory-confirmed cases of SARS-CoV-2 infections and over 4,000 deaths were reported in Zambia (1). Lusaka, the national capital, reported the highest number of cases (77%), with 61% of cases reported among men and more than one-third (32%) of cases involving adults aged 30-44 years (2). Elderly patients (aged >60 years) and those with comorbidities (e.g., HIV, chronic lung disease, hypertension, diabetes, cardiovascular disease, and chronic kidney disease) are at high risk of severe infection and mortality (3,4). Immunosuppression has also been associated with more severe outcomes (5).

HIV remains one of the biggest public health problems globally, with approximately 38.4 million people living with HIV (PLHIV) globally in 2021 and over 650 000 deaths from AIDS-related illnesses in 2021 (6). The HIV epidemic continues to impact a large portion of Zambian adults, with an HIV prevalence of 9.9 (95% CI: 9.1-10.7) among adults aged 15-59 years in 2021 [4].

Current evidence shows that due to aberrant humoral and T-cell-mediated immune responses, PLWH have a higher susceptibility to other infections, such as respiratory infections (7). As more PLHIV age (8), the prevalence of age-related comorbidities among them increases along with the associated risk of severe COVID-19 outcomes. PLHIV with COVID-19 may have more severe clinical presentations and outcomes. The increased risk of SARS-CoV-2 infection or severe disease among PLHIV may be due to immune dysregulation because of CD8-T lymphocyte cell exhaustion (9). T-cell exhaustion occurs secondary to persistently high levels of HIV viral antigen, which leads to strong proinflammatory immune activation and compromised T-cell homeostasis (7). In addition to this HIV- induced immune dysregulation, coronaviruses have been shown to cause transient immune deficiencies that could further weaken the immune system in an already immunocompromised host (10–12). On the other hand, HIV-associated immunosuppression could help temper the cytokine storm of COVID-19 and thus protect against severe forms of COVID-19 (13). In addition, PLHIV on combined antiretroviral treatment (cART) might have a modified risk of infection and a different clinical course of COVID-19 than those not on treatment, as some antiretrovirals may have theoretical activity against SARS-CoV-2 (14–16).

Studies conducted early in the pandemic in the United States of America and Europe showed no difference in the clinical presentation, hospital course and outcomes of COVID-19 between PLHIV and HIV-negative adults (17–20). However, newer evidence from a systematic review and meta-analysis (21), several cohort studies (22–24) and case series (25–27) have shown that PLHIV have a higher risk of severe COVID-19 (hospitalisation, admission to the intensive care unit (ICU), intubation or mechanical ventilation, or death). These studies included PLHIV with suppressed viral loads and different demographic characteristics and comorbidities compared to PLHIV in Africa (9,28,29). The systematic review and meta-analysis conducted by Ssentongo et al in 2021 comprising 22 studies that included over 20 million participants across North America, Africa, Europe, and Asia established that PLHIV had a 24% greater risk of SARS-CoV-2 infection (1.24, 95% [CI 1.05-1.46]) than HIV-negative individuals (21). However, the only African country included in this review was South Africa, one of the most developed economies on the continent. In this meta-analysis, ART (tenofovir and protease inhibitors) did not reduce the risk of SARS-CoV-2 infection or death from COVID-19 among PLHIV (21). In this meta-analysis, most respondents had controlled HIV infection (median CD4 count was 538 cells/μL, and 80% of the PLHIV patients were virally suppressed [than 50 copies of HIV/mL]), and over 96% of PLHIV were on ART(21). In South Africa, a seroprevalence survey among both PLHIV and HIV-negative antenatal women showed that PLHIV had a higher SARS-CoV-2 seroprevalence than antenatal HIV-negative women (30). A prospective cohort study conducted in South Africa showed that PLHIV who were not virally suppressed were more likely to develop symptomatic illness when infected with SAR-CoV-2 (OR 3.3 [95% CI 1.3–8.4]) and shed SARS-CoV-2 for longer (hazard ratio 0.4 [95% CI 0.3–0.6]) compared with HIV-uninfected individuals (31).

Several seroprevalence surveys of SARS-CoV-2 antibody and HIV testing in the general population have shown that PLHIV are less likely to be seropositive than HIV-uninfected individuals (32–35). In these surveys, poor HIV control (evidenced by either high viral load, low CD4 count or not yet on ART) was associated with a poorer antibody response to SARS-CoV-2 infection (32–35). A cross-sectional SARS-CoV-2 seroprevalence survey conducted in Zambia found no association between SARS-CoV-2 antibody seroprevalence and HIV status (36). As this study was limited to only Kabwe District, the findings of this study may not be generalizable to the rest of the country. The lower seroprevalence rates among PLHIV could be due to HIV-induced impaired humoral response, as HIV targets CD4+ T lymphocytes. CD4+ T cells are pivotal in orchestrating both the humoral and cellular immune responses to vaccination and prior infections and have an important role in antibody production (37).

In sub-Saharan Africa, including Zambia, there is a growing body of evidence on coinfection between HIV and COVID-19 with regional differences. Moreover, Zambia has a large population of PLHIV, whose demographic composition and comorbidity profile are different from most of the populations on which the current literature on COVID-19 in PLHIV is based. The aim of this study was to compare the SARS-CoV-2 test positivity, clinical presentation, and predictors of test-specific SARS-CoV-2 positivity between PLHIV and people who are HIV negative in six districts of Zambia. Information is needed to inform policy and interventions focusing on reducing COVID-19 morbidity among PLHIV.

## Materials and Methods

### Study design

We used secondary de-identified data from three different SARS-CoV-2 prevalence surveys conducted in six districts during the upswing of Zambia’s first COVID-19 wave in July 2020. These datasets were made available upon request from the MOH on the 21^st^ of December 2021.

### Study setting

The study was conducted in six out of the 116 districts of Zambia (Lusaka, Ndola, Livingstone, Kabwe, Nakonde and Solwezi) across six provinces (Lusaka, Copperbelt, Southern, Central, Muchinga and North-western) of Zambia. These districts were purposively selected due to high SARS-CoV-2 prevalence, being major gateways or transport hubs and to ensure adequate urban rural disaggregation.

### Study participants

Participants in the three surveys included community members from the selected districts, who were drawn from a cross-sectional cluster-sample survey of households in the six districts of Zambia. Within each district, 16 standardised enumeration areas were randomly selected as primary sampling units using probability proportional to size. All households within each standardised enumeration area were listed, and 20 households from each standardised enumeration area were selected using simple random sampling: people seeking care at outpatient departments (OPDs) in the selected health facilities within these districts and healthcare workers (HCWs) in these same facilities.

### Inclusion criteria

Only individuals (of any age) who had slept in the house the night before the survey was done were eligible to participate in the household survey. In the outpatient survey, participants were randomly selected from all the patients attending 20 conveniently selected outpatient clinics at hospitals and health centres in the six districts. HCWs were selected from the same 20 health facilities (HFs) conveniently selected in the outpatient survey in the six districts. A convenience sample of HCWs at the selected HFs who were present during the survey dates were recruited. For smaller HFs (e.g., health centres), all HCWs were included; for larger HFs (e.g., hospitals), 50 HCWs were invited to participate.

### Sampling technique and sample size estimation

As our study used secondary data, there was no a priori sample size calculation. Participants could accept either PCR or ELISA or both, and we analysed the outcomes of the two different tests separately. For the analysis of overall SARS-CoV-2 PCR test positivity and seropositivity, we used all respondents who took the respective test modality. For the comparison of symptoms reported at enrolment (during survey administration?) Once infected with SARS-CoV-2, we only used the subset of respondents who had a positive PCR test. For the determination of factors associated with SARS-CoV-2 positivity among PLHIV, we used all the PLHIV who accepted either test modality (Figure 1).

To determine if the sample used in our analysis for predictors of SARS-CoV-2 positivity had enough statistical power to make an inference, the sample size by test type was calculated using the following formula.

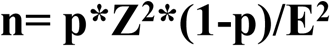

where

n is the required sample size

P is the percentage occurrence of a state or condition E is the percentage maximum error required

Z is the value corresponding to the level of confidence required

With 4642 respondents who were tested by PCR only, the minimum sample size needed for the study to have 80% power is 259; the study therefore has enough power to make statistically significant inferences.

With 4717 respondents who were tested by ELISA only, the minimum sample size needed for the study to have 80% power is 76; the study therefore has enough power to make statistically significant inferences.

### Variables

#### Outcome variable

The outcome variable was SARS-CoV-2 positivity. Participants could be tested by PCR, ELISA, or both. We analysed the two different outcomes separately (PCR and ELISA). PCR and ELISA results were used to determine test-specific positivity and factors associated with being SARS-CoV-2 positive. For the analysis of symptomology, we only used the subset of respondents who had a positive PCR test.

#### Independent variables

The independent variables were demographic characteristics (age, sex) and self-reported medical history (tuberculosis, malaria, diabetes, cardiac disease, hypertension, asthma, chronic obstructive pulmonary disease, chronic kidney disease, liver disease, cancer, any other past medical history). SARS-CoV-2 exposure was defined as any history in the one month prior to testing of household contact with a confirmed case of COVID-19, social contact with a confirmed case of COVID-19, domestic or international travel, in-person attendance to workplace/school/daycare, visit to a health care facility, number of visits to market/grocer and usual means of transportation.

#### Data collection procedures

Data on independent variables were collected through administration of a standardised questionnaire by trained personnel that included information about demographics, self-reported medical history including symptomatology prior to testing, and SARS-CoV-2 exposure history. Next, the specimen for the PCR test was collected by using nasopharyngeal swabs (1 per participant) collected into a cryovial with viral transport medium for detection of SARS-CoV-2 RNA using real-time reverse transcription PCR (38). Additionally, blood samples were collected by finger prick using the BD microtainer EDTA cryovial tube system for detection of anti-spike protein IgG SARS-CoV-2 antibodies using Euroimmun ELISA (PerkinElmer, Waltham, MA, USA) in single replicate according to the manufacturer’s instructions (39).

#### Data management and analysis

All statistical analyses were performed using R statistical software (40). Data from the three surveys were merged for analysis. We then assessed for completeness and missingness by running frequency tables and cross-tabulations of all variables. Due attention was given to skip patterns before using multiple imputations using chained equations using the MICE package in R statistical software to impute values for variables with equal to or less than five percent missing values. Variables with greater than five percent missingness were dropped from subsequent analysis. We then performed diagnostic analyses to identify sample distribution and outliers to guide subsequent statistical tests. The continuous variable age was recoded into a categorical variable age group. A logical variable “any symptom” was recoded from the variable symptom, with two levels, “true or false”. If true, the respondent presented with at least one symptom. Similarly, another logical variable, “any comorbid”, was recoded from those respondents who had reported at least one comorbid medical condition excluding HIV, which was analysed separately.

SARS-CoV-2 positivity was calculated as the number of positive results divided by the total number of tests performed (for a given test modality). PCR and ELISA positivity estimates were reported separately. Descriptive statistics were used to summarise participant characteristics. Stratified analysis was used to compare self-reported HIV status-specific positivity rates. The Shapiro‒Wilk test was used to determine the distribution of continuous variables. Pearson chi-square and Fisher’s exact tests were used to determine associations between the independent and outcome variables. Wilcoxon rank sum tests were used to determine the difference in the medians for continuous variables between PLHIV and HIV-negative participants.

Multilevel logistic regression was used to determine the factors associated with SARS-CoV-2 positivity among PLWH by PCR and ELISA as well as the strength of association. To account for the spatial clustering of participants in each of the six districts, we assumed a random intercept; that is, each of the 6 districts had its unique intercept. In contrast, we assumed fixed effects for all other covariates (that is, an estimated mean effect for the 6 districts). Variables controlled for included age, ART use, sex, visit to a health facility, in-person attendance at a workplace, school or day care and the presence of any comorbid medical condition. Investigator-led stepwise elimination was used to derive the final model using the AIC and BIC values for each test modality until all predictors had a p value of 0.5 or less. Adjusted odds ratios and their 95% confidence intervals (CIs) (p<0.05) were computed.

#### Ethical considerations

The study was conducted under the public health response authority from the Ministry of Health; therefore, permission from the Permanent Secretary, Technical Services at the Ministry of Health was obtained to access the data currently held at the MOH. The study used secondary data and did not involve any interaction with human subjects. The data were deidentified and kept secure. No names of participants were obtained. The data set was handled with confidentiality and only used for the purposes of this study. The data were not subject to undue prejudice. Ethical approval to conduct the study was obtained from the ERES converge ethical review board (**Ref. No 2021-November-010),** and research authority was obtained from the National Health Research Authority (**NHRA-316/06/10/2022**).

## Results

### Participants

A summary of the recruitment algorithm of study participants from the different primary studies is shown in Figure 1A>. A total of 7,328 respondents were approached in all three studies, with most respondents from the household survey (64%) (Figure 1A). A total of 7,092 (96.6%) participants agreed to participate in the respective surveys: 4469 (95.3%) completed the household survey, 1,952 (98.3%) completed the OPD survey and 660 (99.5%) completed the HCW survey (Figure 1A). A total of 5938 (83.7%) of the participants approached consented to SARS-CoV-2 testing. With 4717 (79.4%) tested by ELISA, 4642 (78.2%) tested by PCR, and 3421 (57.6%) tested by both test modalities (Figure 1B), we did not analyse those tested by both test modalities separately.

**Figure 1A:**
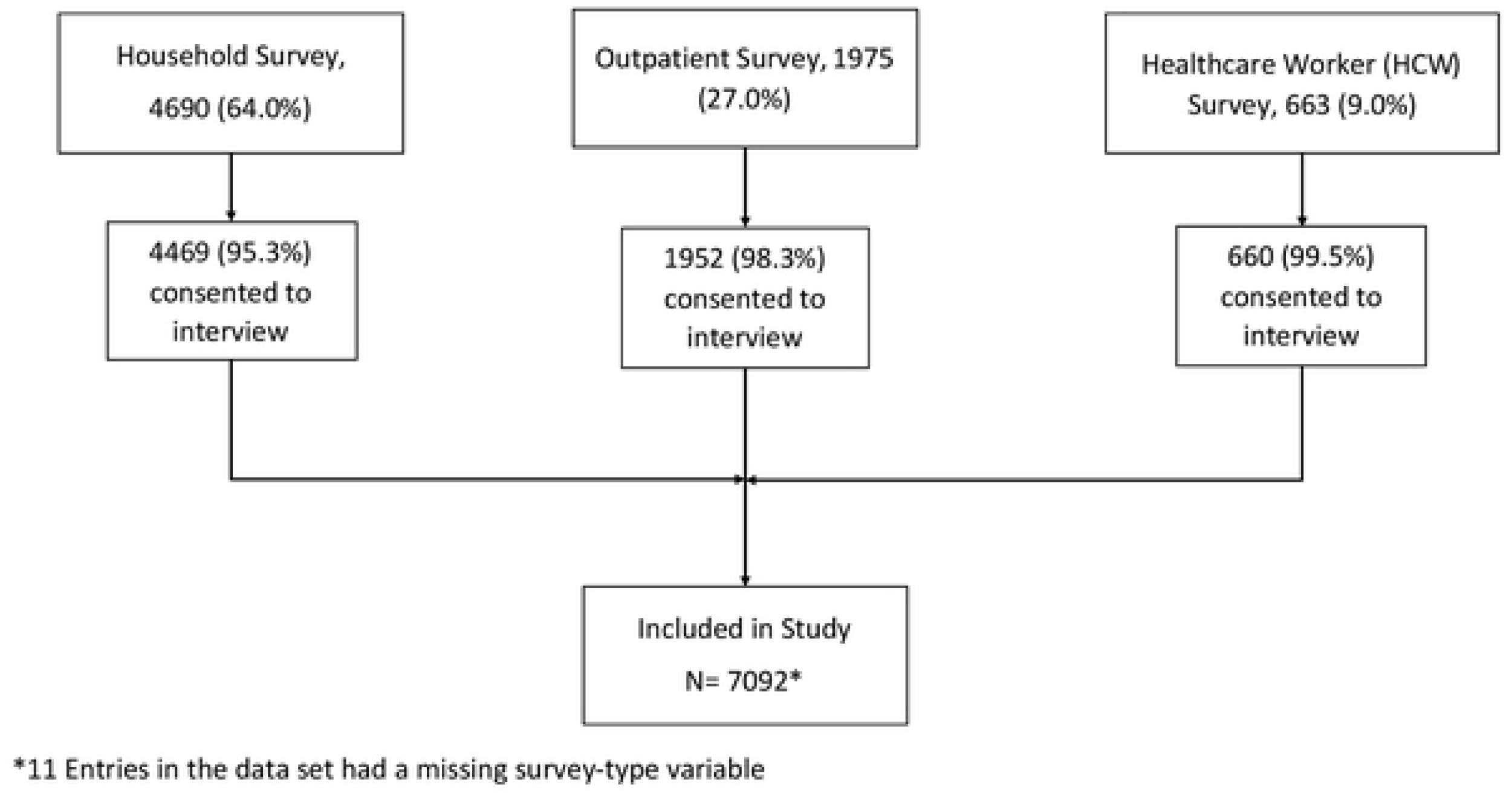
Summary of the study participant recruitment algorithm by study type.

**Figure 1B:**
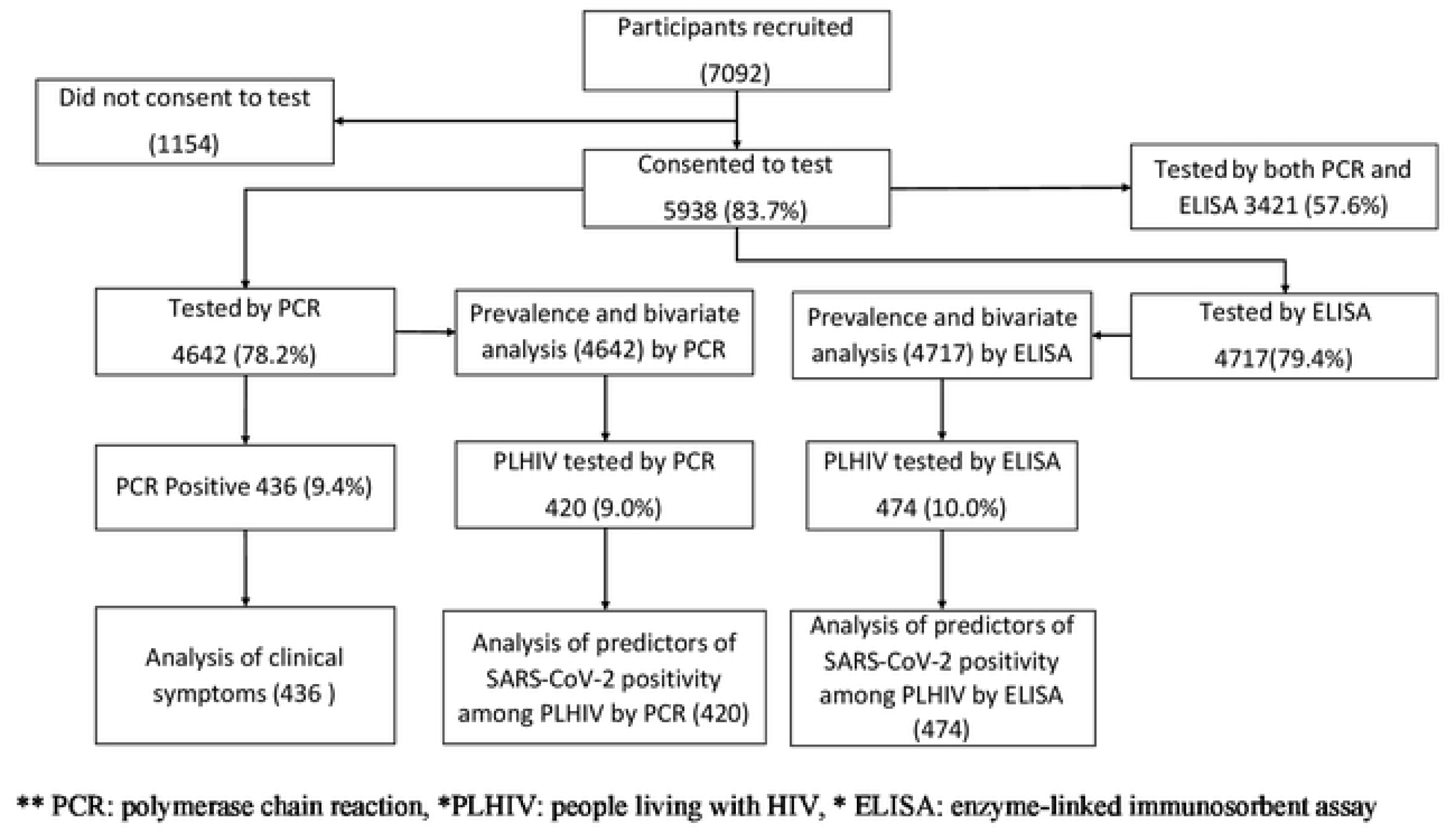
Outline of the algorithm of the selection of study participants into the different arms of the study.

### Demographic characteristics of study participants

The median age among all participants who consented to take part in the study was 28.0 years (IQR: 19.0-41.0). More female participants (61.6%) were enrolled, and most participants had attended secondary education (37.9%) and did not have any comorbid medical conditions (82.0%). In the entire sample, the HIV prevalence was 8.8 percent, and among those who were HIV positive, only 2.4 percent were not on ART (Table 1). Most respondents came from Lusaka district (23.8%), followed by Solwezi district (18.2%), with the least number of respondents from Nakonde district (10.6%) (Table 1). The demographic characteristics of respondents who were tested by either PCR or ELISA were similar to those from the overall sample (Table 1). The demographic characteristics of those tested by PCR or ELISA were similar (Table 1). Compared to HIV-negative study participants, PLHIV were older (median ages: 42 years versus 27 years, p value < 0.001), and a greater proportion had at least one comorbidity other than HIV (44.5% versus 16.7%, p value < 0.001) (Table 2).

**Table 1.** Demographic characteristics of study participants tested for SARS-CoV-2 infection, 6 districts, Zambia, July 2020.

| **Variable** | **Overall (N= 7092)**<br>**n (%)** | **Tested by**<br>**ELISA<sup>1</sup>**<br>**(N=4717), n (%)** | **Tested by PCR**<br>**(N=4642), n**<br>**(%)** |
| --- | --- | --- | --- |
| **Sex** |  |  |  |
| Male | 2716 (38.4) | 1786 (37.9) | 1854 (39.9) |
| Female | 4352 (61.6) | 2931 (62.1) | 2788 (60.1) |
| Missing | 24 |  |  |
| **Age, years** |  |  |  |
| median<br>(IQR<sup>3</sup>) | 28.0 [19.0;41.0] | 29.0 [21.0;42.0] | 28.0 [19.0;41.0] |
| **Age group,**<br>**years** |  |  |  |
| <19 | 1842 (26.1) | 1009 (21.4) | 1231 (26.5) |
| 20-29 | 1926 (27.3) | 1378 (29.2) | 1224 (26.4) |
| 30-39 | 1395 (19.8) | 984 (20.9) | 919 (19.8) |
| 40-49 | 847 (12.0) | 593 (12.6) | 549 (11.8) |
| 50-59 | 548 (7.8) | 397 (8.4) | 374 (8.1) |
| >60 | 495 (7.0) | 356 (7.5) | 345 (7.4) |
| Missing | 39 |  |  |
| **Education** |  |  |  |
| Primary | 2221 (32.0) | 1393 (29.5) | 1399 (30.1) |
| Secondary | 2632 (37.9) | 1863 (39.5) | 1816 (39.1) |
| Higher | 1608 (23.2) | 1231 (26.1) | 1111 (23.9) |
| None | 485 (7.0) | 230 (4.9) | 316 (6.8) |
| Missing | 146 |  |  |
| **Any**<br>**Comorbidity** |  |  |  |
| No | 5815 (82.0) | 3806 (80.7) | 3751 (80.8) |
| Yes | 1277 (18.0) | 911 (19.3) | 891 (19.2) |
| **HIV** |  |  |  |
| No | 5853 (91.2) | 4243 (90.0) | 4222 (91.0) |
| Yes | 565 (8.8) | 474 (10.0) | 420 (9.0) |
| Missing | 674 |  |  |
| **On ART** |  |  |  |
| Yes | 537 (97.6) | 407 (97.8) | 365 (97.9) |
| No | 13 (2.4) | 9 (2.2) | 8 (2.1) |
| NA | 6542 | 4301 | 4269 |
| **Survey** |  |  |  |
| Outpatient | 1952 (27.6) | 1576 (33.4) | 1395 (30.1) |
| Household | 4469 (63.1) | 2616 (55.5) | 2823 (60.8) |
| Healthcare worker | 660 (9.3) | 525 (11.1) | 424 (9.1) |
| Missing | 11 |  |  |
| **District** |  |  |  |
| Lusaka | 1690 (23.8) | 1152 (24.4) | 977 (21.0) |
| Livingstone | 1134 (16.0) | 836 (17.7) | 781 (16.8) |
| Nakonde | 753 (10.6) | 499 (10.6) | 308 (6.6) |
| Ndola | 1234 (17.4) | 979 (20.8) | 975 (21.0) |
| Kabwe | 987 (13.9) | 582 (12.3) | 657 (14.2) |
| Solwezi | 1294 (18.2) | 669 (14.2) | 944 (20.3) |
| 1 PCR: polymerase chain reaction, 2 ELISA: -enzyme-linked immunosorbent assay, 3 Combination of either ELISA and PCR positive and ELISA or PCR negative, 4 HCW: healthcare worker. |  |  |  |

**Table 2.** Demographic characteristics of respondents stratified by HIV status and SARS-CoV-2 test type.

|  | Tested by PCR |  |  | Tested by ELISA |  |  |
| --- | --- | --- | --- | --- | --- | --- |
|  | HIV status |  |  | HIV status |  |  |
|  | Negative<br>(N=4222)<br>n (%) | Positive<br>(N=420)<br>n (%) | p value | Negative<br>(N=4254)<br>n (%) | Positive<br>(N=463)<br>n (%) | p value |
| **Sex<sup>1</sup>** |  |  | 0.0004 |  |  | 0.0004 |
| Male | 1720 (40.7) | 134 (31.9) |  | 1646 (38.7) | 140 (30.2) |  |
| Female | 2502 (59.3) | 286 (68.1) |  | 2608 (61.3) | 323 (69.8) |  |
| **Age<sup>2</sup>** |  |  | <0.0001 |  |  | <0.0001 |
| Median (IQR) | 27.0<br>[18.0;39.0] | 42.0<br>[34.0;51.0] |  | 28.0<br>[20.0;40.0] | 42.0<br>[35.0;50.0] |  |
| **Age group<sup>1</sup>** |  |  | <0.0001 |  |  | <0.0001 |
| <19 | 1221 (28.9) | 10 (2.4) |  | 1001 (23.5) | 8 (1.7) |  |
| 20-29 | 1166 (27.6) | 58 (13.8) |  | 1319 (31.0) | 59 (12.7) |  |
| 30-39 | 812 (19.2) | 107 (25.5) |  | 859 (20.2) | 125 (27.0) |  |
| 40-49 | 422 (10.0) | 127 (30.2) |  | 448 (10.5) | 145 (31.3) |  |
| 50-59 | 305 (7.2) | 69 (16.4) |  | 321 (7.5) | 76 (16.4) |  |
| >60 | 296 (7.0) | 49 (11.7) |  | 306 (7.2) | 50 (10.8) |  |
| **Education completed<sup>1</sup>** |  |  | 0.0021 |  |  | 0.0003 |
| Primary | 1281 (30.3) | 118 (28.1) |  | 1245 (29.3) | 148 (32.0) |  |
| Secondary | 1620 (38.4) | 196 (46.7) |  | 1652 (38.8) | 211 (45.6) |  |
| Higher | 1021 (24.2) | 90 (21.4) |  | 1147 (27.0) | 84 (18.1) |  |
| None | 300 (7.1) | 16 (3.8) |  | 210 (4.9) | 20 (4.3) |  |
| **Any comorbidities<sup>1</sup>** |  |  | <0.0001 |  |  | <0.0001 |
| No | 3518 (83.3) | 233 (55.5) |  | 3529 (83.0) | 277 (59.8) |  |
| Yes | 704 (16.7) | 187 (44.5) |  | 725 (17.0) | 186 (40.2) |  |
| **PCR result<sup>1</sup>** |  |  | 0.0277 |  |  | 0.0311 |
| Negative | 3838 (90.9) | 368 (87.6) |  | 4086 (96.1) | 454 (98.1) |  |
| Positive | 384 (9.1) | 52 (12.4) |  | 168 (3.9) | 9 (1.9) |  |
| **On ART** |  |  | NA |  |  | NA |
| Yes |  | 365 (97.9) |  |  | 407 (97.8) |  |
| No |  | 8 (2.1) |  |  | 9 (2.2) |  |
| Missing |  | 47 |  |  | 47 |  |
| **Survey<sup>1</sup>** |  |  | <0.0001 |  |  | <0.0001 |
| Outpatient | 1202 (28.5) | 193 (46.0) |  | 1354 (31.8) | 222 (47.9) |  |
| Household | 2621 (62.1) | 202 (48.1) |  | 2405 (56.5) | 211 (45.6) |  |
| Healthcare worker | 399 (9.5) | 25 (6.0) |  | 495 (11.6) | 30 (6.5) |  |
| **District<sup>1</sup>** |  |  | <0.0001 |  |  | <0.0001 |
| Lusaka | 903 (21.4) | 74 (17.6) |  | 1040 (24.4) | 112 (24.2) |  |
| Livingstone | 699 (16.6) | 82 (19.5) |  | 745 (17.5) | 91 (19.7) |  |
| Nakonde | 289 (6.8) | 19 (4.5) |  | 471 (11.1) | 28 (6.0) |  |
| Ndola | 868 (20.6) | 107 (25.5) |  | 872 (20.5) | 107 (23.1) |  |
| Kabwe | 552 (13.1) | 105 (25.0) |  | 490 (11.5) | 92 (19.9) |  |
| Solwezi | 911 (21.6) | 33 (7.9) |  | 636 (15.0) | 33 (7.1) |  |
| <sup>1</sup> Pearson-Chi-square, <sup>2</sup>Wilcoxon rank sum test |  |  |  |  |  |  |

### SARS-CoV-2 positivity

Among all participants, SARS-CoV-2 positivity was 9.4% and 3.8% by PCR and ELISA, respectively. SARS-CoV-2 positivity by PCR was 6.6%, 13.4% and 7.8% in the HCW survey, outpatient survey and household survey, respectively. SARS-CoV-2 positivity by ELISA was 2.3%, 5.4% and 3.1% in the HCW survey, outpatient survey and household survey, respectively (Table 2). There was no difference in positivity by sex of the respondent (OR 1.2, [PCR], OR 1.1, [ELISA]) (Table 2). Among those for whom PCR testing was completed, positivity differed significantly by age: compared to respondents aged ≤19 years, respondents within the age groups 20 to 29 and 30 to 39 years had higher odds (OR 1.2, 95% CI: 1.12-1.96 and OR 1.4, 95% CI: 1.06-1.93, respectively) of testing SARS-CoV-2 positive by PCR (Table 2).

PLHIV had increased odds of testing positive for SARS-CoV-2 by PCR (OR 1.4, CI: 1.03-1.91) but had reduced odds of an ELISA-positive test (OR 0.5, 95% CI: 0.23-0.90). Among PLHIV, those not on ART had increased odds of SARS-CoV-2 infection by PCR (OR: 7.5, 95% CI: 1.72-32.7)) (Table 2).

There was district-by-district variation in SARS-CoV-2 test positivity. Livingstone had the highest test positivity (13.8%) by PCR, while Nakonde had the highest test positivity by ELISA (9.6%) (Table 2). Compared to people who had not received any education, people who had completed secondary and higher education had higher odds of SARS-CoV-2 infection (OR 1.9, 95% CI: 1.19-3.26, OR 1.9, 95% CI: 1.18-3.33, respectively). Excluding HIV as a comorbidity, there was no difference in SARS-CoV-2 test positivity between people who had comorbid medical conditions and those who did not (OR 0.9, 95% CI: 0.71,1.20) (Table 2).

### Predictors of SAR-CoV-2 positivity among PLHIV

In the multilevel logistic regression model, PLHIV not receiving ART was an independent predictor of SARS-CoV-2 positivity by PCR (aOR 5.78, 95% CI: 1.35-24.76). All the other covariates were insignificant, and none of the covariates were significant in the ELISA analysis (Table 3). There was a similar finding in the ordinary logistic regression model (Table 4).

**Table 3.** Prevalence and bivariate analysis of SARS-CoV-2 positivity by test modality in six districts of Zambia, July 2020.

|  | Tested by PCR (N=4642) * |  | Tested by ELISA (N=4717) * |  |
| --- | --- | --- | --- | --- |
|  | Prevalence n (%) | OR (95%CI) | Prevalence n (%) | OR (95%CI) |
| **Overall** | 436 (9.4) |  | 177(3.8) |  |
| **Sex** |  |  |  |  |
| Female | 247(8.9) | Ref | 105(3.6) | Ref |
| Male | 189(10.2) | 1.2(0.96-1.42) | 72(4.0) | 1.1(0.83-1.53) |
| **Age group** |  |  |  |  |
| <19 | 93(7.6) | Ref | 34(3.4) | Ref |
| 20-29 | 132(10.8) | 1.5(1.12-1.96) | 46(3.3) | 1.0(0.63-1.56) |
| 30-39 | 96(10.4) | 1.4(1.06-1.93) | 41(4.2) | 1.3(0.79-1.99) |
| 40-49 | 55(10.0) | 1.4(0.96-1.93) | 24(4.0) | 1.2(0.70-2.05) |
| 50-59 | 28(7.5) | 1.0(0.63-1.52) | 16(4.0) | 1.2(0.64-2.17) |
| >60 | 32(9.3) | 1.3(0.81-1.89) | 16(4.5) | 1.4(0.72-2.44) |
| **Education completed** |  |  |  |  |
| None | 114(8.1) | Ref | 7(3.0) | Ref |
| Primary | 188(10.4) | 1.5(0.9-2.53) | 50(3.6) | 1.2(0.57-2.90) |
| Secondary | 116(10.4) | 1.9(1.19-3.26) | 83(4.5) | 1.5(0.73-3.57) |
| Higher | 18(5.7) | 1.9(1.18-3.33) | 37(3.0) | 1.0(0.46-2.44) |
| **Survey** |  |  |  |  |
| Healthcare worker | 28(6.6) | Ref | 12(2.3) | Ref |
| Outpatient | 187(13.4) | 2.2(1.47-3.37) | 85(5.4) | 2.4(1.37-4.73) |
| Household | 221(7.8) | 1.2(0.81-1.84) | 80(3.1) | 1.4(0.76-2.62) |
| **District** |  |  |  |  |
| Kabwe | 30(4.6) | Ref | 14(2.4) | Ref |
| Lusaka | 106(10.8) | 2.5(1.70-3.93) | 53(4.6) | 2.0(1.11-3.69) |
| Livingstone | 108(13.8) | 3.4(2.24-5.18) | 20(2.4) | 1.0(0.50-2.03) |
| Nakonde | 17(5.5) | 1.2(0.65-2.22) | 48(9.6) | 4.3(2.41-8.22) |
| Ndola | 106(10.9) | 2.6(1.70-3.93) | 26(2.7) | 1.1(0.58-2.19) |
| Solwezi | 69(7.3) | 1.7(1.07-2.59) | 16(2.4) | 1.0(0.48-2.08) |
| **Comorbid medical condition** |  |  |  |  |
| No | 354(9.4) | Ref | 133(3.5) | Ref |
| Yes | 82(9.2) | 0.9(0.71-1.20) | 44(4.8) | 1.4(0.98-1.97) |
| **HIV Status** |  |  |  |  |
| Negative | 384(9.1) | Ref | 168(3.9) | Ref |
| Positive | 52(12.4) | 1.4(1.03-1.91) | 9(1.9) | 0.5(0.23-0.90) |
| **On ART** |  |  |  |  |
| Yes | 43(11.8) | Ref | 9(2.2) | Ref |
| No | 4(50.0) | 7.5(1.72-32.7) | 0(0) | 0.0 |
| Missing | 359 |  | 168 |  |
| **Household contact COVID** |  |  |  |  |
| No | 432(9.4) | Ref | 176(3.8) | Ref |
| Yes | 4(12.5) | 1.4(0.41-3.54) | 1(3.2) | 0.9(0.05-4.02) |
| **Travel** |  |  |  |  |
| None | 348(9.1) | Ref | 152(3.9) | Ref |
| International | 5(16.7) | 2.0(0.68-4.87) | 2(6.9) | 1.8(0.29-6.20) |
| Domestic | 83(10.8) | 1.2(0.94-1.56) | 23(3.0) | 0.8(0.48-1.17) |
| **Visit Health Facility** |  |  |  |  |
| No | 375(10.2) | Ref | 143(3.9) | Ref |
| Yes | 61(6.3) | 0.6(0.44-0.78) | 34(3.3) | 0.8(0.57-1.22) |
| **In-person<sup>1</sup>** |  |  |  |  |
| No | 313(9.0) | Ref | 127(3.6) | Ref |
| Yes | 123(10.6) | 1.2(0.96-1.48) | 50(4.1) | 1.2(0.82-1.60) |
| **Transport** |  |  |  |  |
| Walking | 251(9.3) | Ref | 97(3.6) | Ref |
| Personal car | 45(10.7) | 1.2(0.83-1.62) | 18(4.0) | 1.1(0.64-1.80) |
| Taxi | 45(11.1) | 1.2(0.86-1.68) | 13(3.5) | 1.0(0.51-1.67) |
| Bike | 9(6.6) | 0.7(0.32-1.29) | 7(5.6) | 1.6(0.65-3.24) |
| Minibus | 86(8.8) | 0.9(0.73-1.22) | 42(4.0) | 1.1(0.76-1.58) |

**Table 4.** Multilevel multivariate analysis of predictors of SARS-CoV-2 positivity among PLHIV by test modality in six districts of Zambia, July 2020.

| **Predictors** | **PCR Positivity (420)**<br>**aOR (95%CI)** | **ELISA positivity (474)**<br>**aOR (95%CI)** |
| --- | --- | --- |
| **Sex** |  |  |
| Male | 1.32(0.70-2.52) | 1.62(0.41-6.47) |
| **Any comorbid** |  |  |
| Yes | 1.34(0.72-2.49) | 3.72(0.94-14.68) |
| **On ART** |  |  |
| No | **5.78(1.35-24.76)** | 5.35(0.54-53.42) |
| **HH contact COVID** |  |  |
| Yes | 6.39(0.29-142.79) | NA |
| **In person** |  |  |
| Yes | 1.32(0.66-2.62) | 0.71(0.13-3.84) |
| **Visit Health facility** |  |  |
| Yes | NA | 1.92(0.54-6.85) |
| **Age** | NA | 0.97(0.92-1.03) |

PCR: polymerase chain reaction
ELISA: enzyme-linked immunosorbent assay
aOR: adjusted odds ratio
95%CI: 95%: confidence interval
ART: antiretroviral therapy
In-person attendance at workplace, school or day-care
HH household contact
NA Variables not included in the final model of the respective analysis.
We assumed a random intercept for district and survey type; that is, each of the 6 districts and survey had its unique intercept. In contrast, we assumed fixed effects for all other covariates (that is, an estimated mean effect for the 6 districts). Variables controlled for included age, ART use, sex, visit to a health facility, in-person attendance to a workplace, school, or day care and for the presence of any comorbid medical condition).

**Table 5:** Predictors of SARS-CoV-2 test positivity by test type, multivariate logistic regression.

|  | **PCR Positivity**<br>**(420)**<br>**aOR(95%CI)** | **ELISA positivity**<br>**(474)**<br>**aOR(95%CI)** |
| --- | --- | --- |
| **Sex** |  |  |
| Male | 1.24(0.65-2.34) | 1.58(0.36-6.40) |
| **Age** |  | 0.97(0.91-1.02) |
| **Any Comorbid** |  |  |
| Yes | 1.32(0.71-2.48) | 4.5(1.14-20.37) |
| **On ART** |  |  |
| No | **5.24(1.19-22.22)** | 4.89(0.23-37.94) |
| **HH Contact COVID** |  |  |
| Yes | 9.79(0.28-371.89) | NA |
| **In Person** |  |  |
| Yes | 1.33(0.66-2.59) | 0.87(0.18-3.17) |
| **District** |  |  |
| Kabwe | Ref |  |
| Lusaka | 6.49(1.85-31.05) | 5.41(0.70-112.35) |
| Livingstone | 9.11(2.86-41.36) | NA |
| Nakonde | 2.3(0.11-19.77) | 5.98(0.22-162.36) |
| Ndola | 7.2(2.25-32.89) | 5.36(0.77-107.99) |
| Solwezi | 2.55(0.32-16.72) | NA |
| **Visit HF** |  |  |
| Yes | NA | 2.08(0.55-7.52) |

PCR: polymerase chain reaction
ELISA: enzyme-linked immunosorbent assay
adjusted odds ratio
95%: confidence interval
ART: antiretroviral therapy
In-person attendance at workplace, school or day-care
HH household contact
NA Variables not included in the final model of the respective analysis.
Variables controlled for included age, ART use, sex, visit to a health facility, in-person attendance to a workplace, school, or day care and for the presence of any comorbid medical condition.

**Table 6:** Comparison of symptoms reported by SARS-CoV-2 PCR-positive respondents at enrolment by self-reported HIV status in six districts of Zambia, July 2020.

| Symptom | PCR<sup>1</sup> positive (N=436) |  |  |
| --- | --- | --- | --- |
|  | Prevalence, n (%) | OR<sup>2</sup> | 95% CI<sup>3</sup> |
| **Any symptom** |  |  |  |
| HIV-negative | 98(25.4) | Ref | — |
| PLHIV<sup>4</sup> | 22(44.0) | **2.31** | **1.25, 4.21** |
| **Fever** |  |  |  |
| HIV-negative | 21(5.4) | Ref | — |
| PLHIV | 3(6.0) | 1.11 | 0.26, 3.38 |
| **Chills** |  |  |  |
| HIV-negative | 13(3.4) | Ref | — |
| PLHIV | 3(6.0) | 1.83 | 0.41, 5.94 |
| **Cough** |  |  |  |
| HIV-negative | 19(4.9) | Ref | — |
| PLHIV | 8(16.0) | **3.68** | **1.44, 8.67** |
| **SOB<sup>5</sup>** |  |  |  |
| HIV-negative | 1(0.3) | Ref | — |
| PLHIV | 2(4.0) | **16** | **1.51, 349** |
| **Fatigue** |  |  |  |
| HIV-negative | 11(2.8) | Ref | — |
| PLHIV | 2(4.0) | 1.42 | 0.22, 5.50 |
| **Anorexia** |  |  |  |
| HIV-negative | 2(0.5) | Ref | — |
| PLHIV | 4(8.0) | **16.7** | **3.17, 123** |
| **Myalgia** |  |  |  |
| HIV-negative | 9(2.3) | Ref | — |
| PLHIV | 1(2.0) | 0.85 | 0.05, 4.69 |
| **Arthralgias** |  |  |  |
| HIV-negative | 8(2.1) | Ref | — |
| PLHIV | 1(2.0) | 0.96 | 0.05, 5.42 |
| **Rhinorrea** |  |  |  |
| HIV-negative | 19(4.9) | Ref | — |
| PLHIV | 7(14.0) | **3.14** | **1.17, 7.62** |
| **Wheezing** |  |  |  |
| HIV-negative | 1(0.3) | Ref | — |
| PLHIV | 1(2.0) | 7.86 | 0.31, 201 |
| **Altered taste** |  |  |  |
| HIV-negative | 3(0.8) | Ref | — |
| PLHIV | 1(2.0) | 2.61 | 0.13, 20.8 |
| **Abdominal pain** |  |  |  |
| HIV-negative | 10(2.6) | Ref | — |
| PLHIV | 5(10.0) | **4.18** | **1.26, 12.3** |
| **Diarrhoea** |  |  |  |
| HIV-negative | 3(0.8) | Ref | — |
| PLHIV | 2(4.0) | 5.32 | 0.69, 32.9 |
| **Chest pain** |  |  |  |
| HIV-negative | 9(2.3) | Ref | — |
| PLHIV | 2(4.0) | 1.75 | 0.26, 7.02 |
| **Headache** |  |  |  |
| HIV-negative | 44(11.4) | Ref | — |
| PLHIV | 5(10.0) | 0.86 | 0.29, 2.11 |
| **Any other<sup>6</sup>** |  |  |  |
| HIV-negative | 15(3.9) | Ref | — |
| PLHIV | 5(10.0) | 2.75 | 0.86, 7.48 |

<sup>1</sup>PCR: polymerase chain reaction
<sup>2</sup> OR: odds ratio
<sup>3</sup>95% confidence interval
<sup>4</sup>PLHIV: people living with HIV.
<sup>5</sup> SOB: shortness of breath.
<sup>6</sup>Any other symptom not listed

### Clinical features of SAR-CoV-2-positive cases

Among participants who tested positive for SARS-CoV-2 by PCR, most were asymptomatic at the time of testing (74.6% among the HIV negative and 56.0% among PLHIV). However, PLHIV had increased odds of being symptomatic compared to HIV-negative participants (OR 2.3, 95% CI 1.3-4.2). PLHIV also more frequently presented with cough, shortness of breath, anorexia, rhinorrhea, and abdominal pain compared to HIV-negative SARS-CoV-2 respondents (Table 4).

## Discussion

The aim of this study was to compare the SARS-CoV-2 test positivity, clinical presentation, and predictors of SARS-CoV-2 positivity between PLHIV and people who are HIV negative in six districts of Zambia. Our findings showed that among respondents to surveys administered during the early COVID-19 pandemic, PLHIV had higher odds of testing positive for SARS-CoV-2 by PCR than HIV-negative respondents, and at the time of testing for SARS-CoV-2 infection, PLHIV who tested PCR-positive were more frequently symptomatic than participants who were HIV negative. Among PLHIV, not being on ART was an independent predictor of SARS-CoV-2 PCR positivity.

PLHIV had higher odds of testing positive for SARS-CoV-2 by PCR than HIV-negative respondents. In this study, PLHIV were older and had more comorbidities than the HIV-negative participants; this could have affected our findings, as both increasing age and a greater number of comorbidities have been shown to increase the risk of SARS-CoV-2 infection (3–5) Additionally, early in the first wave, PLHIV may have had an additional risk of exposure to SARS-CoV-2 due to more frequent hospital visits for medical review and drug collection. This finding is consistent with findings from a large, pooled analysis of 22 studies from across the world from early 2020 through 2021 that showed that PLHIV had 24% higher odds of SARS-CoV-2 infection than their HIV-negative counterparts (30,41,42). Underlying socioeconomic disparities in PLHIV have been shown to increase exposure to SARS-CoV-2 (43). Among PLHIV, not being on ART was an independent predictor of SARS-CoV-2 PCR positivity. Most studies performed on HIV and SARS-CoV-2 coinfection were inconclusive about the effect of ART, as most PLHIV in these studies were on suppressive ART (42). PLHIV who have not achieved viral suppression through ART may have a compromised immune system that leaves them vulnerable to opportunistic infections (44).

In the bivariate analysis, there were reduced odds of a positive ELISA test among PLHIV compared to the HIV-negative respondents. As ELISA detects the body’s humoral response to SARS-CoV-2 infection, factors that affect the body’s immune system may affect the reliability of this test. HIV affects CD4+ T cells, which play a key role in orchestrating the immune system (9). As such, PLHIV may have a delayed or decreased response in producing antibodies to SARS-CoV-2 after a natural infection. Several seroprevalence surveys of SARS-CoV-2 antibody and HIV testing have shown that PLHIV are less likely to be seropositive than HIV-uninfected individuals (45–47). In these surveys, high HIV viral load, low host CD4 count or not yet on ART was associated with poorer antibody response to SARS-CoV-2 infection (44,46,47). Further research may be required to verify this finding among PLHIV in Zambia.

Overall, PLHIV were more likely to present with a symptomatic SARS-CoV-2 infection at the time of testing. Additionally, cough, shortness of breath, anorexia, rhinorrhea, and abdominal pain were more likely in PLHIV. These findings suggest that at the time of enrolment, PLHIV were more likely to have a symptomatic SARS-CoV-2 infection than HIV-negative people. Our findings are consistent with a review of 22 studies that included over 20 million participants across North America, Africa, Europe, and Asia that showed an increased risk of severe outcomes in PLHIV (31,48), although most PLHIV in these studies had been on suppressive ART. A study by the WHO showed that PLHIV were at increased risk of severe or critical disease at hospital admission (aOR 1.1, 95% CI 1.0–1.1) compared to HIV-negative individuals (48).

Our study had several limitations. The timing of the study at the beginning of the first wave in Zambia could have biased positive results to those respondents more linked to international or domestic travel. A past medical history of underlying conditions such as HIV was self-reported. As we merged data from three surveys, some study-specific variables had much missingness in the combined data set and therefore could not be analysed further. This resulted in fewer explanatory variables in our analysis. Additionally, PCR positivity can be long-lasting, especially among people who are immunocompromised. As such, people who are PCR positive may be weeks away from the initial infection.

Despite these limitations, we believe that our findings are valid due to the large sample size, inclusion of study participants across different sociodemographic characteristics, and geographic settings and due to consistency in the findings of this study with other local and international studies.

### Conclusion

These findings suggest that at the time of enrolment in the study, during the first COVID-19 wave in Zambia, PLHIV were at an increased risk of SARS-CoV-2 infection, as evidenced by PCR, but were less likely to have had a prior infection, as evidenced by ELISA. Among the infected individuals, PLHIV were more likely to be symptomatic. Among PLHIV, not being on ART was independently associated with an increased risk of SARS-CoV-2 infection. Interventions focusing on ensuring early access to COVID-19 vaccinations, testing and ART might reduce COVID-19 morbidity among PLHIV.

## Data Availability

The data that support the findings of this study are available from the Ministry of Health of the Government of Zambia, but restrictions apply to the availability of these data, which were used under permission for the current study, and so are not publicly available. However, the data are available from the authors upon reasonable request and with permission of the Permanent Secretary of the Ministry of Health.

## Acknowledgements

We thank the Ministry of Health, Zambia, for providing SARS-CoV-2 prevalence data in Zambia.

## Author contributions

Stephan Longa Chanda was the principal investigator.

Stephen Longa Chanda, Cephas Sialubanje, Mukumbuta Nawa, Nyambe Sinyange, Warren Malambo, James Zulu, Dabwitso Banda1, Paul Zulu1, Lloyd B. Mulenga and Jonas Hines designed the study.

Warren Malambo, Nyambe Sinyange, James Zulu, Dabwitso Banda, and Jonas Hines collected the data.

Stephan Chanda, and Jonas Z Hines analysed the data.

Stephen Longa Chanda, Cephas Sialubanje, Mukumbuta Nawa, Nyambe Sinyange, Warren Malambo, James Zulu, Dabwitso Banda, Paul Zulu, Lloyd B. Mulenga, Jonas Hines wrote the manuscript.

